# Investigation of BRCA1 exon 11 genetic variations in breast cancer among Libyan women

**DOI:** 10.1101/2023.10.13.23296599

**Authors:** Halla Elshwekh, Inas Alhudiri, Hamza Musbah Abdul Jalil, Abdenaser A. Mohamed, Ahmed Elkikli, Naji Jornaz, Nabil Enattah

**Affiliations:** Genetic engineering department, Libyan Biotechnology Research Center-Tripoli-Libya; Tripoli University Hospital, Department of Oncology, Tripoli, Libya

**Keywords:** Keyword: genetic variants, breast cancer, unclassified variants, Libya

## Abstract

**Introduction:** The majority of hereditary breast and ovarian cancers are associated with mutations in two genes, breast cancer type 1 and 2 susceptibility genes (BRCA1 and BRCA2). Here, we describe for the first time analysis of BRCA1 exon 11 in 48 Libyan breast cancer patients with a family history of cancer.

**Methods:** All patients had a family history of cancer and were included in the study only if they have a family history of breast or ovarian cancer, male breast cancer, or triple negative tumors. PCR was performed using specific primer pairs spanning BRCA1 exon 11 followed by Sanger sequencing. Genetic analysis was done using Sequencher® 5.1.

**Results:** We identified 12 genetic variants in BRCA1 exon 11. Three variants were novel (c.1019T>C, c.2363T>G, and c.3192T>C). c.2363T>G was predicted by SIFT as damaging. Six variants were of unknown significance (c.918T>C, c.1853G>C, c.1886G>A, c.2215A>G, c.2612C>T, c.3113A>C and c.3784T>C), and 3 were classified as benign in ClinVar database (c.918T>C, c.2082C>T and c.2311T>C).

**Conclusions:** Scanning of the entire BRCA1 is needed to identify any associated deleterious mutations. Although the clinical importance of unclassified variants is unknown, the association of certain variants with deleterious mutations and contralateral breast cancer warrants genetic testing and counseling in the Libyan population.

## Introduction

Despite recent advances in the early diagnosis and treatment of breast cancer, it remains an important cause of cancer mortality in women, and it accounts for about 23% of cancers among women worldwide (Bray *et al*, 2018). Breast cancer is not only the most common malignancy in women throughout the world, accounting for 22.9% of cancer in women, it is also one of the major causes of death (Bray *et al*, 2018). In the Middle East, it was reported as the leading tumour in women in all cancer registries, accounting for 27.7-38.2% of all reported tumours. According to the Middle East Cancer Consortium, the age-standardized incidence rate (ASRs) per 100,000 women was higher among Israeli Jews (93.1), similar to the rates reported in North America and Western Europe (Uhrhammer *et al*, 2008).

The most common cancer of women in Libya is breast cancer (20.6%) (Elzouki *et al*, 2018). Most Libyan patients present with advanced disease and are often younger than in Europe, in line with the pattern common in North Africa (Boder *et al*, 2011). The incidence of breast cancer in women is highest after age 50 years, but 5-12% of this cancer develops in women under 45 years (Juwle & Saranath, 2012).

About 5-10% of breast cancers have a hereditary predisposition (Al Hannan *et al*, 2019). Hereditary breast cancer is characterized by early onset, high incidence of bilateral disease, and repetitive correlation with ovarian cancer (Olopade et al. 2008).

Understanding the risk factors for breast cancer enables identification of women at increased risk and implementation of risk-modifying interventions, both individually and population wise (Shah *et al*, 2018). Although a family history of breast and/or ovarian cancer is common in women diagnosed with breast or ovarian cancer, less than 10% of all breast cancers and less than 15% of ovarian cancers are associated with germline (inherited) genetic mutations (Antoniou *et al*, 2011; Cao *et al*, 2016). The most common cause of hereditary breast and ovarian cancers is the hereditary breast and ovarian cancer syndrome (HBOC), which is associated with germline mutations in BRCA1 and BRCA2. It has been estimated that the HBOC syndrome accounts for about 5% of breast cancer cases ((Nik-Zainal *et al*, 2016).

Over 1000 different mutations in BRCA1 and BRCA2 have been reported. Mutations occur throughout the entire coding region, but most of them lead to truncation of the translated protein. Despite the wide range of mutations, founder effects have led to higher prevalence of some mutations in certain geographic or ethnic populations (Boeri *et al*, 2011; Michailidou *et al*, 2015).

The largest exon in human BRCA1 is exon 11, which consists of 3426 bases. By splicing, it produces three different transcripts of mature BRCA1 protein (the full isoform, D11 isoform, and D11 q isoform). Splicing regulatory sequences are located at the 5′ end of exon 11, and important sequences at its 3′ end. In particular, a strong splicing enhancer was found adjacent to the downstream 5′ splice site. By using minigene and deletion analyses, it was shown to minimize competition from an upstream 5′ splice site and so ensures long exon inclusion (Raponi *et al*, 2014; El Khachibi *et al*, 2015).

The identification of sequences and proteins relevant for the regulation of BRCA 1 exon 11 has provided better knowledge on how this exon is recognized and may represent an important step toward understanding how large exons are regulated (Cherbal *et al*, 2012b; Raponi *et al*, 2014).

Defining women at high risk enables implementation of preventive strategies and early diagnosis of the disease. Here, we describe for the first time analysis of BRCA1 exon 11 in 48 Libyan breast cancer patients with a family history of cancer.

## Martials and Methods

### Study population

The patients were recruited from the oncology departments of the main public hospitals in Tripoli (Tripoli Central Hospital and Tripoli Medical Center), which provide oncology services to most of the region. Blood samples were collected from the patients in EDTA-containing Vacutainer tubes by trained members of the hospital staff and stored at −20°C until DNA extraction.

The laboratory analyses were performed at the genetic engineering department of the Biotechnology Research Center in Tripoli. The study was conducted on 48 Libyan women attending the oncology department of Tripoli Central Hospital or Tripoli Medical Center. To be included in the study, the patient had to be a female aged 18-60 years fulfilling the following criteria: Breast cancer at young age (<40), Family history of any cancer, cancer in both breasts in the same woman, Both breast and ovarian cancers in either the same woman or the same family, Multiple breast cancers, Two or more primary types of BRCA1- or BRCA2-related cancers in a single family member, Cases of male breast cancer or Triple negative breast cancer. These criteria were defined with reference to the criteria for consideration of *BRCA1/2* genetic testing developed by the National Comprehensive Cancer Network.

Ethical approval was granted by the Bioethics Committee of the Biotechnology Research Centre (Reference No. BEC-BTRC 03-2014; Annex I). Volunteering patients and healthy controls signed informed consent forms.

The participants filled out a self-administered questionnaire on age, origin, residence region, milk intake and problems associated with it, presence of abdominal symptoms, smoking status, diabetes, hypertension, cancer, helicobacter infection, cardiac disease, and osteoporosis.

### Laboratory analyses

DNA was extracted from peripheral blood using the QIAamp DNA Blood Mini kit (Qiagen). The concentration of the purified DNA was estimated by using a Nanodrop Lite spectrophotometer (Thermo Scientific) and its integrity was checked by agarose gel electrophoresis.

PCR was carried out in a volume of 50 µl containing 10 µl of 5x GoTaq™ reaction buffer, 2 µl of template, 1.5 mM MgCl_2_, 1 µl of dNTPs, 1.5 µl of forward primer, 1.5µl of reverse primer, and 0.25 μl (0.8 U) of GoTaq™ DNA polymerase. Preparation of the PCR tubes was carried out on ice.

The PCR was started with denaturation at 95°C for 2 min. This was followed by 25 cycles of denaturation at 95°C for 35 sec, annealing at 53°C for 30 sec, and extension at 72°C for 35 sec. Then, one cycle of extension at 72°C and samples were stored at 4°C until sequencing.

The PCR products were purified by using a QIAquick Purification Kit™ to remove excess salts, PCR primers, and dNTPs. The purified PCR products were cycle-sequenced using BigDye® Terminator v3.1 Cycle Sequencing kit as manufacturer’s protocol. Extension products purification was performed by using BigDye® XTerminator™. Capillary electrophoresis was performed on an Applied Biosystems (3500xL Advanced Genetic Analyzer). The run was set up using Data Collection Software. Sequencing data were analysed

Using Sequencher Software. Analysis protocols, including base caller and mobility file, were applied, the analysis was run, and the data were reviewed.

### Statistical analysis

All statistical analyses were performed using SPSS for Windows, Version 24.0 (SPSS Inc., Chicago, Illinios, USA). Pearson’s X² test was used. A p value < 0.05 was considered to be statistically significant for all analyses.

## Results and discussion

### Clinical and pathological characteristics of sample population

A total of 48 samples from patients with breast cancer were investigated for *BRCA1* exon 11 genetic variations. The median age at diagnosis was 46 years (mean ± SD = 45 ± 15.9). Twenty one patients had a family history of breast cancer, 16 of them in first degree relatives and the rest in second degree relatives. All the patients had unilateral breast cancer.

Most patients were < 50 years, but about one-third of them were diagnosed at a much younger age (≤ 40). Most patients were married and only 16.7% were holding university degrees. Most of the patients were either obese or overweight (∼68%) and 31% of them were obese, making them at higher risk of breast cancer.

Table 1 summarizes the age at diagnosis, marital status and educational level of the breast cancer patients.

**Table 1.** Demographics of study population.

| Parameter | N (48) | % |
| --- | --- | --- |
| Age |  |  |
| Mean Age | 47.15 | - |
| mean±SD | 11.35 | - |
| Age at diagnosis |  |  |
| Age at onset $\leq 40$ | 16 | 33 |
| Age at onset 40-50 | 13 | 27 |
| Age at onset $>50$ | 19 | 39.5 |
| Marital status: |  |  |
| Married | 41 | 85.4 |
| Divorced | 1 | 2 |
| Single | 5 | 10.4 |
| Widow | 1 | 2 |
| Education |  |  |
| No education | 20 | 41.7 |
| Diploma | 5 | 10.4 |
| High school | 11 | 22.9 |
| Preparatory | 2 | 4.2 |
| Primary | 2 | 4.2 |
| University | 8 | 16.7 |

**Table 2.** Clinical characteristics.

| Parameter | N | % |
| --- | --- | --- |
| BMI |  |  |
| **Healthy 18.5-24.9** | 7 | 14.6 |
| **Obese $\geq 30$** | 15 | 31.3 |
| **Overweight 25-29.9** | 18 | 37.5 |
| **Underweight $<18.5$** | 5 | 10.4 |
| Hx of benign breast lump |  |  |
| **No** | 31 | 64.6 |
| **Yes** | 17 | 35.4 |
| Current treatment |  |  |
| **Chemotherapy** | 38 | 79 |
| **Radiotherapy** | 21 | 44 |
| **Hormonal therapy** | 14 | 29 |
| Surgery type |  |  |
| **Lumpectomy** | 2 | 4.2 |
| **Mastectomy** | 37 | 77 |
| **None** | 9 | 18.8 |

Sixty-four percent of the patients did not report any previous history of benign breast lump. Also, the majority of patients has breastfed their babies (83%) and were parous (77%). Half of the patients had more than three children and 29.2% had more than five. Most patients had their first live birth before age 30. Between 81 and 85% of them reported no history of diabetes mellitus or hypertension and almost two-thirds of them (64%) had no accompanying heart disease (Table 3). Clinical and reproductive data are listed in table 5.

**Table 3.** Presence of comorbidities in study patients.

| Disease | No of patients | % |
| --- | --- | --- |
| Diabetes |  |  |
| **No** | 39 | 81.3 |
| **Yes** | 9 | 18.8 |
| Hypertension |  |  |
| **No** | 41 | 85.4 |
| **Yes** | 7 | 14.6 |
| History of heart disease |  |  |
| **No** | 31 | 64.6 |
| **Yes** | 17 | 35.4 |

**Table 4.**
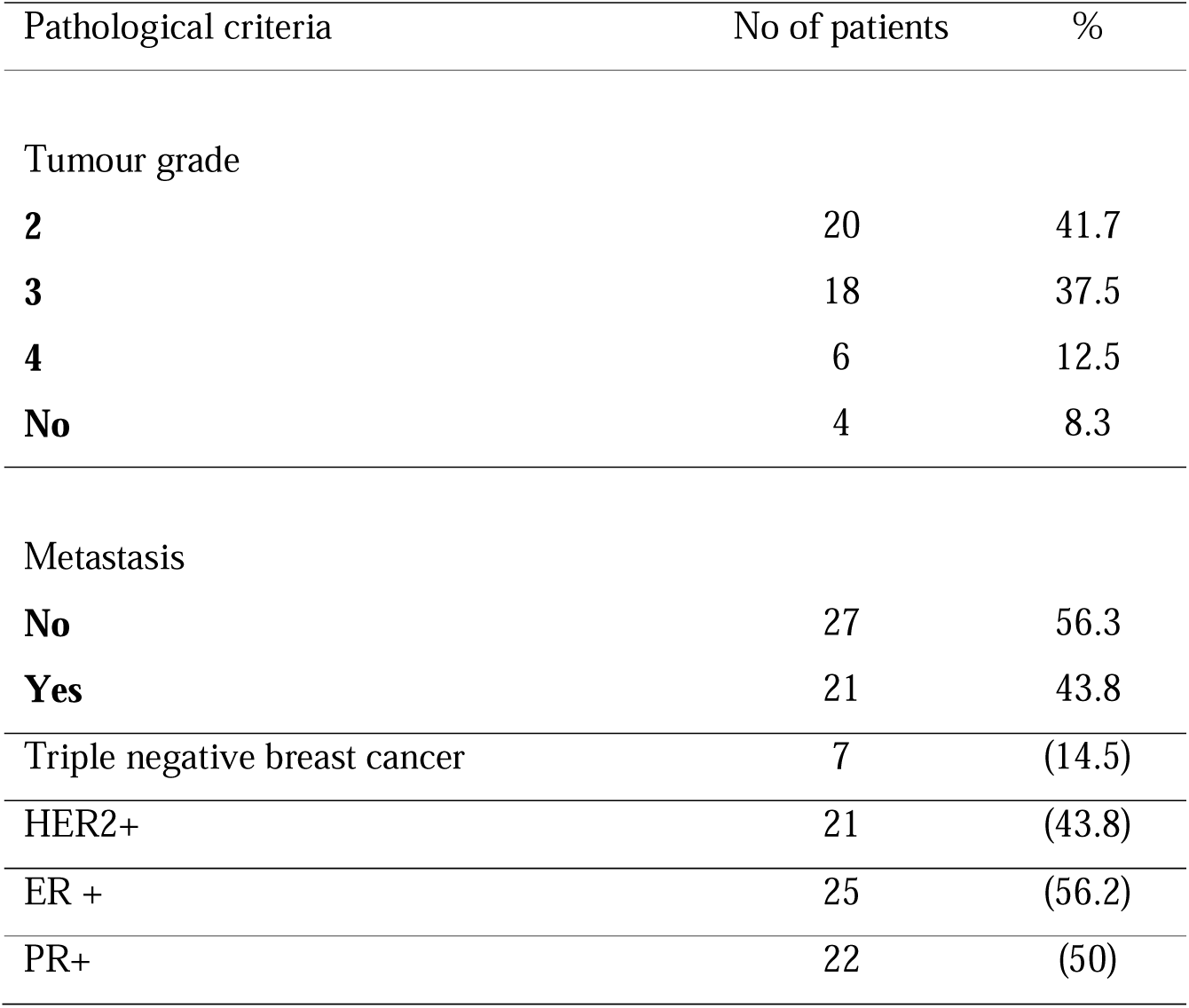
Pathological characteristics of study patients.

| Pathological criteria | No of patients | % |
| --- | --- | --- |
| Tumour grade |  |  |
| **2** | 20 | 41.7 |
| **3** | 18 | 37.5 |
| **4** | 6 | 12.5 |
| **No** | 4 | 8.3 |
| Metastasis |  |  |
| **No** | 27 | 56.3 |
| **Yes** | 21 | 43.8 |
| Triple negative breast cancer | 7 | (14.5) |
| HER2+ | 21 | (43.8) |
| ER + | 25 | (56.2) |
| PR+ | 22 | (50) |

**Table 5.** Reproductive characteristics of study patients.

| Parameter | n | % |
| --- | --- | --- |
| Breastfeeding |  |  |
| **No** | 8 | 16.7 |
| **Yes** | 40 | 83.3 |
| Age at menarche (years) |  |  |
| **7-11** | 9 | 18.8 |
| **12-13** | 22 | 45.8 |
| **>13** | 17 | 35.4 |
| Age at menopause (years) |  |  |
| **≤40** | 6 | 12.5 |
| **>40** | 42 | 87.5 |
| Parity |  |  |
| **Yes** | 11 | 77.1 |
| Parity (By no of children) |  |  |
| **0** | 11 | 22.9 |
| **1-3** | 13 | 27.1 |
| **>3** | 24 | 50 |
| Parity (by no of pregnancies) |  |  |
| **>5 full-time pregnancies** | 14 | 29.16 |
| Age at first live birth |  |  |
| **No birth** | 11 | 22.9 |
| **<20 years old** | 9 | 18.8 |
| **20-24 years old** | 7 | 14.6 |
| **25-29 years old** | 15 | 31.3 |
| Family history of breast cancer |  |  |
| **First degree relative** | 21 | 41.6 |
| **Second degree relative** | 16 |  |
| Family history of cancer | 5 | 58.3 |
| Clomid/other assisted fertilization | 28 | 27 |
| **Clomid** | 13 | 4.2 |
| **Similar** | 2 |  |
| Oral contraceptives use |  |  |
| **No** | 27 | 56.3 |
| **Yes** | 21 | 44 |

Table 4 shows that most tumors were mildly or moderately differentiated cancers and a few were poorly differentiated. Distant metastasis was observed in 43.8% of cases. As for hormonal receptor status, more than half of the tumors were ER positive (56%) and 50% were PR positive. Surprisingly, 43.8% of the cases were HER2 positive. Seven out of 48 (14.5%) patients were negative for these markers.

### PCR amplification of *BRCA1* exon 11 fragments

Extracted genomic DNA was amplified by PCR using the primer sequences shown in supplementary table 1. *BRCA1* exon 11 was divided into five parts named A, B, C, D, and E for PCR before Sanger sequencing. Figures 1 shows PCR amplification of BRCA1 exon 11A 1% TBE agarose gel electrophoresis. Lanes 1-18 are amplified BRCA1 exon 11 A DNA at a size of 816 bp. M =100 bp DNA ladder used to determine the approximate size of the PCR products.

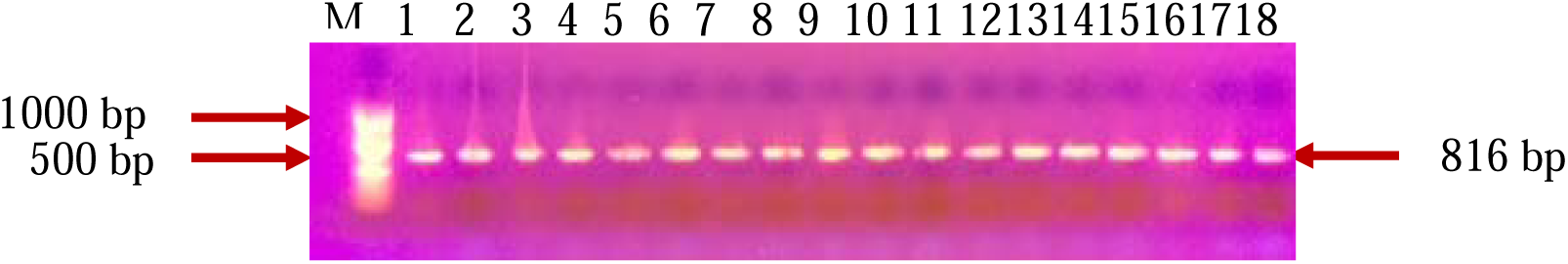

### Genotyping of BRCA1 exon 11

There is a tremendous amount of information on the effect of germline variations in BRCA1 and BRCA2 genetic predisposition to breast cancer in different populations. However, to our knowledge, this is the first report on genetic variants of BRCA1 exon 11 in Libyan breast cancer patients. In this study, 48 Libyan breast cancer patients with a family history of breast cancer and fulfilling the other eligibility criteria who were admitted to the main hospitals in Tripoli were screened for BRCA1 exon 11 germline genetic variations using Sanger sequencing.

Successful sequences were obtained for 33 patients. Unfortunately, no sample was successfully screened for the whole length of exon 11 of the BRCA1 gene. This study identified 12 BRCA1 exon 11 genetic variants in 33 patients with a family history of cancer (Tables 6 and 7). Nine variants were previously reported in NCBI SNP database and ClinVar and three novel variants. Most of the reported variants are of unknown significance (6/9) and three were classified according to ClinVar database as benign or likely benign. The novel variants have not been described in any of the databases (NCBI db SNP, ClinVar, ExAc, 1000 genome) or reported in studies conducted in Arab countries. Sequences for the three novel variants were uploaded into Genbank as accession numbers MW716256, MW716257, MW716258, MW716259, and MW716260.

**Table 6.** Polymorphisms and genetic variants in *BRCA1* exon 11 in Libyan breast cancer patients.

| HGVS name | AA change | dbSNP | Clin Var | SIFT prediction | Variation type | Genotype Frequency n (%) |  |
| --- | --- | --- | --- | --- | --- | --- | --- |
| NM_007294.3:c.918T>C | p.Asn306= | rs1555592835 | Likely benign | Tolerated | synonymous | TT | 6 (85.7) |
|  |  |  |  |  |  | TC | 0 |
|  |  |  |  |  |  | CC | 1 (12.2) |
| NM_007294.3:c.1853G>C | p.Arg618Thr | rs876659527 | Uncertain significance | Damaging | Missense variant | GG | 11/14 (79) |
|  |  |  |  |  |  | CG | 3/14 (21.4) |
|  |  |  |  |  |  | CC | 0 |
| NM_007294.3:c.1886G>T | p.Arg629Ile | rs876660144 | Uncertain significance | Damaging | Missense variant | AA | 0/14 |
|  |  |  |  |  |  | GG | 0/14 |
|  |  |  |  |  |  | AG | 1/14 (7.14) |
| NM_007294.3:c.2215A>G | p.Lys739Gln | rs56329598 | Uncertain significance | Tolerated | Missense variant | AA | 14/18 (77.7) |
|  |  |  |  |  |  | GG | 0 |
|  |  |  |  |  |  | AG | 4/18 (22.2) |
| NM_007294.3:c.2082C>T | p.Ser694= | rs1799949 | Benign | N/A | synonymous | CC | 12/20 (60) |
|  |  |  |  |  |  | TT | 2/20 (10) |
|  |  |  |  |  |  | CT | 6/20 (30) |
| NM_007294.3:c.2311T>C | p.Leu771= | rs16940 | Benign | Tolerated | synonymous | CT | 9/18 (50) |
|  |  |  |  |  |  | TT | 9/18 (50) |
|  |  |  |  |  |  | CC | 0 |
| NM_007294.3:c.2612C>T | p.Pro871Leu | rs799917 | Uncertain significance | Tolerated | Missense | CC | 4/14 (28.5) |
|  |  |  |  |  |  | TT | 6/14 (42.8) |
|  |  |  |  |  |  | CT | 4/14 (28.5) |
| NM_007294.3:c.3113A>G | p.Glu1038Ala | rs16941 | Uncertain significance | Damaging | Missense | AA | 6/11 (54.5) |
|  |  |  |  |  |  | GG | 0 |
|  |  |  |  |  |  | AG | 5/11 (45.5) |
| NM_007294.3:c.3784T>C | p.Ser1262Pro | rs1011096937 | Uncertain significance | Tolerated | Missense | TT | 17/18 (94.4) |
|  |  |  |  |  |  | CC | 0 |
|  |  |  |  |  |  | CT | 1/18 (5.5) |

**Table 7.** Novel polymorphisms and genetic variants in BRCA1 Exon 11 in Libyan breast cancer patients.

| Variant | Variation Type | AA change | Polyphen-2 | SIFT | Mutation taster | HGVS name | Genotype frequency | N (%) |
| --- | --- | --- | --- | --- | --- | --- | --- | --- |
| c.1019T>C | Missense | p.Val340Ala<br>V340A | Possibly<br>damaging | Tolerated | Protein may be<br>affected | NM_007294.3:r.1251 | TT | 4/6(66.6) |
|  |  |  |  |  | Splice site<br>changes | NM_007294.3:c.1019 | TC | 1/6(16.6) |
|  |  |  |  |  |  |  | CC | 1/6(16.6) |
| c.2363T>G | Missense | p.val788glycine<br>V788G | Benign | Damaging | Protein may be<br>affected | NM_007294.3:r.2595 | TT | 16/18 (88.8) |
|  |  |  |  |  | Splice site<br>changes | NM_007294.3:c.2363 | GG | 2/18 (11.1) |
|  |  |  |  |  |  | NP_009225.1:p.788 | TG |  |
| c.3192T>C | synonymous | p.Ser1064=<br>S1064S | N/A | Tolerated | Unkown effect | NM_007294.3:r.3424 | TT | 10/11 (90.9) |
|  |  |  |  |  |  | NM_007294.3:c.3192 | CC | 0 |
|  |  |  |  |  |  | NP_009225.1:p.1064 | CT | 1/11 (9) |

**Table 8.** Allele frequency of study variants registered in SNP data base.

| SNP | Allele 1 n (%) | Allele 2 n (%) | Allele 3 n (%) |
| --- | --- | --- | --- |
| Rs1555592835 | TT | CC |  |
|  | 5 | 1 |  |
|  | T | C |  |
|  | 10(83.3) | 2(16.6) |  |
| Rs876659527 | GG | CG |  |
|  | 11 | 1 |  |
|  | G | C |  |
|  | 23(95.8) | 1(4.1) |  |
| Rs56329598 | AA | AG |  |
|  | 10 | 1 |  |
|  | A | G |  |
|  | 21(95.4) | 1(4.5) |  |
| Rs8766601 | GG | GA |  |
|  | 10 | 1 |  |
|  | G | A |  |
|  | 21(95.4) | 1(4.5) |  |
| Rs1799949 | CC | CT | TT |
|  | 10 | 4 | 4 |
|  | C | T |  |
|  | 24(66.6) | 12(33.3) |  |
| Rs16940 | TT | CT |  |
|  | 6 | 5 |  |
|  | T | C |  |
|  | 17(77.2) | 5(22.7) |  |
| Rs799917 | TT | CC | CT |
|  | 5 | 3 | 3 |
|  | T | C |  |
|  | 13(59.01) | 9(40.9) |  |
| Rs16941 | AG | AA |  |
|  | 3 | 6 |  |
|  | A | G |  |
|  | 15(83.3) | 3(16.6) |  |
| Rs1011096937 | TT | CT |  |
|  | 14 | 1 |  |
|  | T | C |  |
|  | 29(96.6) | 1(3.3) |  |

**Table 9.**
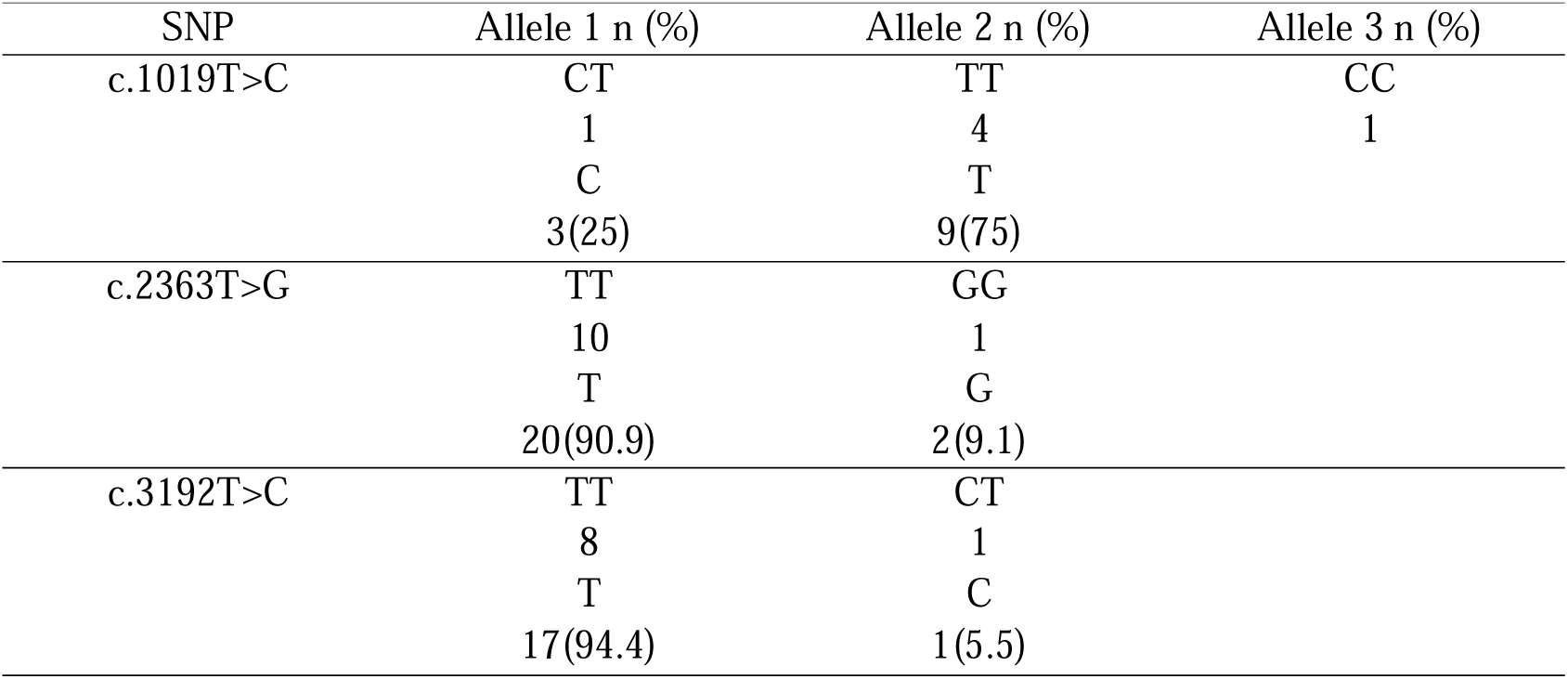
Allele frequency of novel variants in this study.

| SNP | Allele 1 n (%) | Allele 2 n (%) | Allele 3 n (%) |
| --- | --- | --- | --- |
| c.1019T>C | CT<br>1<br>C<br>3(25) | TT<br>4<br>T<br>9(75) | CC<br>1 |
| c.2363T>G | TT<br>10<br>T<br>20(90.9) | GG<br>1<br>G<br>2(9.1) |  |
| c.3192T>C | TT<br>8<br>T<br>17(94.4) | CT<br>1<br>C<br>1(5.5) |  |

The novel c.1019T>C variant was seen in a woman aged >50 years with a family history of prostate cancer but no family history of breast cancer. The variant is predicted to be tolerated by SIFT (Sorting Intolerant From Tolerant is an algorithm that predicts the potential impact of amino acid substitutions on protein function) but possibly damaging in Polyphen-2. The Mutation Taster software classified the variant as a polymorphism with splice site changes and stated that the protein may be affected. MAF for this variant was 25%.

The other new variant, c.2363T>G, was seen in a woman aged >50 years with a family history of breast cancer. SIFT predicts this variant as damaging, and Mutation Taster predicts it as a polymorphism with possible protein effects and splice site changes. Only homozygous genotypes are seen in this variant, TT and GG. MAF (G) is 0.9%

The third novel variant, c.3192T>C, is synonymous and is predicted by Mutation Taster as disease-causing, but it is tolerated by SIFT. This variant was detected in a woman aged >50 years with family history of breast cancer. MAF (C) is 5.5%.

Four of the previously described variants that are considered common genetic variants were detected in this study. Common variants are defined as having a population minor allele frequency > 1% of the population according to 1000 genome data, whereas rare variants have a frequency ≤ 1% population minor allele frequency. These variants (c.2311T>C, c.2612C>T, c.3113A>G and c.2082C>T) have also been reported in Tunisia, Algeria, Morocco and Bahrain (Cherbal *et al*, 2012a; Mahfoudh *et al*, 2012; Tazzite *et al*, 2012; Fourati *et al*, 2014; Al Hannan *et al*, 2019).

Whether these common variants are independently associated with a risk of breast cancer is unknown. A previous study showed that comparison of genotype frequency between cases and controls could detect variants of unknown significance of BRCA genes that cause disease (Figueiredo *et al*, 2011). Of these variants, c.2311T>C (rs16940) SNP seemed to add an increased risk of contralateral breast cancer among women who had known deleterious mutations in BRCA1, but it was associated with a reduced risk in non-BRCA1 and non-BRCA2 carriers (Figueiredo *et al*, 2011). Table 11 shows the presence of the variants detected in Arab women from 5-8 variants as in Libyan women.

**Table 11.** The distribution of SNPs detected in Libyan women compared to other Arab countries.

| Variation ID | SNP | Tunis | Algeria | Morocco | Egypt | Lebanon | Saudi A |
| --- | --- | --- | --- | --- | --- | --- | --- |
| dbSNP |  |  |  |  |  |  |  |
| rs1555592835 | c.918T>C |  |  |  |  |  |  |
| rs876659527 | c.1853G>C |  |  |  |  |  |  |
| Rs56329598 | c.2215A>C |  |  |  |  |  |  |
| rs876660144 | c.1886G>T |  |  |  |  |  |  |
| rs1799949 | c.2082C>T | ✓ | ✓ | ✓ |  |  | x |
| rs16940 | c.2311 | ✓ | ✓ | ✓ |  |  | x |
| rs799917 | c.2612C>T | ✓ | ✓ | ✓ |  |  | x |
| rs16941 | c.3113A>C | ✓ | ✓ | ✓ |  |  | x |
| rs1011096937 | c.3784 |  |  |  |  |  |  |

This study has several limitations, largely due to budget constraints. These include the small sample size and non-inclusion of healthy controls for comparison of the unclassified variants. Nevertheless, other limitations include poor sequencing results of many samples and some samples were not scanned for the whole length of exon 11. It would be advantageous to scan the BRCA1 gene for all possible mutations to draw better conclusions regarding the association of genetic variants with pathogenic mutations. All the patients were specifically asked about their family history of breast cancer, but it is is possible that some of this information is incomplete or inaccurate.

Scanning of the entire BRCA1 is needed to identify any associated deleterious mutations. Although the clinical importance of unclassified variants is unknown, the association of certain variants with deleterious mutations and contralateral breast cancer warrants genetic testing and counseling in the Libyan population.

## Data Availability

All data produced in the present study are available upon reasonable request to the authors

