## supplemental table 1 for "Investigation of BRCA1 exon 11 genetic variations in breast cancer among Libyan women"

| Exon | Forward primer | Reverse primer | Ta (°C) | PCR product (bp) |
| --- | --- | --- | --- | --- |
| Exon 11 A | 5'-atttccacctccaaggtgtatg  30425→30446 | 5'-ttccgataggttttcccaaata  31219←31240 | 58 | 816 |
| Exon 11 B | 5'-tgaaagagttcactccaaatcag  31172→31194 | 5'-ccaggtgcatttgttaacttca  31939←31960 | 59 | 789 |
| Exon 11 C | 5'-atggaaggtaaagaacctgcaa  31845→31866 | 5'-caaaacctagagcctcctttga  32677←32698 | 61 | 854 |
| Exon 11 D | 5'-atatcactgcaggctttcctgt  33381→33402 | 5'-ctctaatttcttggcccctctt  34000←34021 | 61 | 860 |
| Exon 11 E | 5'-gtccagaaaggagagcttagca  33381→33402 | 5'-tgtaaaatgtgctccccaaaag  34000←34021 | 60 | 641 |

**Table 1**: Primer sequences
